# Case fatality risk of novel coronavirus diseases 2019 in China

**DOI:** 10.1101/2020.03.04.20031005

**Authors:** Xiaowei Deng, Juan Yang, Wei Wang, Xiling Wang, Jiaxin Zhou, Zhiyuan Chen, Jing Li, Yinzi Chen, Han Yan, Juanjuan Zhang, Yongli Zhang, Yan Wang, Qi Qiu, Hui Gong, Xianglin Wei, Lili Wang, Kaiyuan Sun, Peng Wu, Marco Ajelli, Benjamin J. Cowling, Cecile Viboud, Hongjie Yu

**Affiliations:** School of Public Health, Fudan University, Key Laboratory of Public Health Safety, Ministry of Education, Shanghai, China; Savaid Medical School, University of Chinese Academy of Sciences, Beijing, China; Division of International Epidemiology and Population Studies, Fogarty International Center, National Institutes of Health, Bethesda, MD, USA; WHO Collaborating Centre for Infectious Disease Epidemiology and Control, School of Public Health, Li Ka Shing Faculty of Medicine, University of Hong Kong, Hong Kong Special Administrative Region, China; Bruno Kessler Foundation, Trento, Italy

**Author notes:** These authors contributed equally to this work (Xiaowei Deng, Juan Yang, Wei Wang, Xiling Wang). Corresponding author to Prof. Hongjie Yu.

## Abstract

**Objective:** The outbreak of novel coronavirus disease 2019 (COVID-19) imposed a substantial health burden in mainland China and remains a global epidemic threat. Our objectives are to assess the case fatality risk (CFR) among COVID-19 patients detected in mainland China, stratified by clinical category and age group.

**Methods:** We collected individual information on laboratory-confirmed COVID-19 cases from publicly available official sources from December 29, 2019 to February 23, 2020. We explored the risk factors associated with mortality. We used methods accounting for right-censoring and survival analyses to estimate the CFR among detected cases.

**Results:** Of 12,863 cases reported outside Hubei, we obtained individual records for 9,651 cases, including 62 deaths and 1,449 discharged cases. The deceased were significantly older than discharged cases (median age: 77 vs 39 years, p<0.001). 58% (36/62) were male. Older age (OR 1.18 per year; 95%CI: 1.14 to 1.22), being male (OR 2.02; 95%CI: 1.02 to 4.03), and being treated in less developed economic regions (e.g., West and Northeast vs. East, OR 3.93; 95%CI: 1.74 to 8.85) were mortality risk factors. The estimated CFR was 0.89-1.24% among all cases. The fatality risk among critical patients was 2-fold higher than that among severe and critical patients, and 24-fold higher than that among moderate, severe and critical patients.

**Conclusions:** Our estimates of CFR based on laboratory-confirmed cases ascertained outside of Hubei suggest that COVID-19 is not as severe as severe acute respiratory syndrome and Middle East respiratory syndrome, but more similar to the mortality risk of 2009 H1N1 influenza pandemic in hospitalized patients. The fatality risk of COVID-19 is higher in males and increases with age. Our study improves the severity assessment of the ongoing epidemic and can inform the COVID-19 outbreak response in China and beyond.

## Introduction

As of March 3, 2020, a total of 80,270 cases of novel coronavirus disease 2019 (COVID-19) have been reported in mainland China, including 2,981 deaths. The outbreak is caused by a novel coronavirus of presumed zoonotic origin, the severe acute respiratory syndrome coronavirus 2 (SARS-CoV-2)^1^. COVID-19 cases have now been identified in 72 countries, some of which have reported onward local transmission and deaths^2^. The unprecedented scale of the epidemic has prompted an urgent need for clinical severity assessment, of which the case fatality risk (CFR) is a key metric.

A few studies have assessed the fatality risk of COVID-19 but estimates have been highly variable. Wu et al. estimated that the fatality risk among hospitalized cases was 14% during the early phase of outbreak in Wuhan^3^. Dorigatti et al. estimated that the CFR among laboratory-confirmed cases was 18% in Hubei province and ranged from 1.2-5.6% outside mainland China^4^. A recent report of the World Health Organization (WHO)-China Joint Mission on Coronavirus Disease 2019 estimated the case fatality risk as 3.8% by dividing the number of deaths at the time of analysis by the number of laboratory-confirmed cases at the time of analysis^5^. They also reported a higher case fatality risk in Hubei than that in other provinces (5.8% vs. 0.7%)^5^. However, those estimates would be a lower bound on the CFR for the laboratory-confirmed cases because many cases were still in the hospital and had not reached a final outcome of either death or discharge after recovery^6^.

In the present study, we aimed to assess the CFR among laboratory-confirmed COVID-19 cases detected in mainland China, stratified by different clinical categories (e.g. mild-, moderate-, severe- and critical-patients) and by age group. We also explored the risk factors associated with fatal outcomes.

## Methods

### Case definitions and surveillance

The National Health Commission of China (NHC) and the Chinese Center for Disease Control and Prevention (China CDC) have launched a new surveillance system to record information on COVID-19 cases since the start of the outbreak of atypical pneumonia cases in Wuhan in late December 2019. A description of the surveillance system is provided elsewhere^7^. As the epidemic evolves, a total of six versions of case definitions for suspected- and laboratory-confirmed-cases have been issued by NHC^7-9^. Details are provided in the Appendix table S1.

Four clinical categories of laboratory-confirmed COVID-19 patients have been identified by NHC, including mild-, moderate-, severe-, and critical-patients ^7-9^. Mild patients, introduced in the fifth and sixth versions of COVID-19 case definition, refer to patients with mild symptoms and no radiographic evidence of pneumonia. Moderate patients, introduced in the fourth version of the case definition, refers to patients with fever, respiratory symptoms, and radiographic evidence of pneumonia. Severe patients, introduced in the second version, refers to patients with any breathing problems, finger oxygen saturation, and low PaO2/FiO2 (PaO2 denotes partial pressure of oxygen in arterial blood; FiO2 denotes fraction of inspired oxygen), etc. Critical patients, a definition used from the very beginning of the outbreak, refer to patients having any respiratory failure, shock, and any other organ failure that requires ICU admission.

Patients were discharged when they met all the following criteria: 1) normal body temperature for more than 3 days, 2) significantly improved respiratory symptoms, 3) significant inflammation absorption in lung radiographic findings, and 4) negative nucleic acid detection by real-time RT-PCR using respiratory specimens on two consecutive days, with a sampling interval ≥1 day^9^.

### Data collection

Daily aggregated data (hereafter called aggregated dataset) on the cumulative number of cases were extracted from the websites of national, provincial, and municipal Health Commissions. Individual records on laboratory-confirmed COVID-19 cases (hereafter called individual dataset) were collected from two official publicly available sources from December 29, 2019 through to February 23, 2020, including: 1) the websites of national, provincial, and municipal Health Commission; 2) the websites of national and local government affiliated medias. Individual information was extracted and entered into a structured database comprising demographic characteristics, dates of symptom onset, first healthcare consultation, hospital admission, official announcement (reporting date), as well as outcome information (e.g. death/discharge and corresponding dates). Each individual record was extracted and entered by three coauthors and was cross-checked to ensure data accuracy. Conflicting information was resolved based on the Health Commission data. Details on the collection of individual data and assessment of completeness of variables used in the study are provided in Appendix Tables S2-3.

### Statistical analysis

We restricted analyses of demographic characteristics, risk factors associated with fatal outcome, and key time to event intervals to the provinces outside Hubei, where the majority of individual records were obtained (97.6%, 9,651/9,886) as of February 23, 2020. We implemented a multivariate logistic regression model to explore the risk factors associated with death. We included age, sex, economic region^10^, time interval from symptom onset to first medical consultation, first hospital admission, and laboratory diagnosis. We categorized China into three economic regions (East, Central, West and Northeast) according to gross domestic product per capita in 2018 (see Appendix Figure S1 for map)^10^.

We estimated key time-to-event distributions including symptom onset to first healthcare consultation, hospital admission, laboratory diagnosis, and death or discharge, and from hospital admission to death or discharge. We fitted three parametric distributions (Weibull, gamma, and lognormal) to time-to-event data and selected the best fit based on the minimum Akaike information criterion.

We used three methods to estimate CFR among COVID-19 cases. Firstly, we calculated a crude CFR based on the cumulative number of deaths divided by the cumulative number of laboratory-confirmed cases, ignoring the time-lag between symptoms onset and death and resulting right-censoring of outcomes^5^.

In a second approach, we adjusted for delays between hospitalization and death to obtain more accurate estimates of CFR, using the method described by Garske et al. for pandemic influenza A/H1N1 in 2009^11^. For above two methods, we used the aggregated dataset as of February 23, and binomial distributions were used to estimate the 95%CIs.

Thirdly, to allow for incomplete information about outcomes, we used survival analyses to allow inclusion of all cases admitted to hospital in the individual dataset, incorporating data for patients who were still in hospital at the time of analysis. In our individual dataset, the outcome was unavailable for some patients because the information was not communicated through public channels, although these patients may have been discharged or died at the time of this writing (hereafter denoted as missing outcome). This is different from the issue of right-censoring for patients still hospitalized whose illnesses have yet to be resolved. The cases who were still hospitalized and those with missing outcome were treated as unresolved in our analysis. A multiple imputation was used to generate outcomes for these patients.

For each date t, we calculated the number of discharged/deceased patients that required imputation by subtracting the number of discharged/deceased patients in our individual dataset from that in the aggregated dataset (Table S4 in Appendix). All these patients with missing data for outcomes before date t were considered for imputation on date t. They were randomly selected as discharge or death according to probability calculated using the density of interval from hospital admission to discharge/death. This imputation procedure was repeated 100 times to generate 100 imputed datasets for further estimation of CFR. We employed a dual-outcome (discharge or death) time to event framework to estimate CFR based on the fraction F1/(F1+F2)^12^. F1 and F2 stand for the admission to death distribution and the admission to discharge distribution, respectively. 

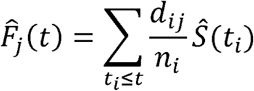

Where, *t*_l_ <…< *t*_*k*_ denotes the distinct observed event times for outcomes (discharge or death), with *d*_*ij*_ representing the number of outcome j that occur at time *t*_*i*_. *Ŝ* is the Kaplan-Meier estimator of overall survival function (combined event of discharge or death)^12^. Then we implemented a 1,000 times bootstraps for estimation of 95%CIs, and used Rubin’s formula to pool all estimates across 100 imputed datasets^13^.

For the survival analysis, we restricted analyses to the provinces outside Hubei, considering the completeness of individual records obtained. When estimating CFR, we excluded cases hospitalized beyond 17 days on each date in the baseline analysis. The choice of 17 days was based on the 90^th^ percentile of the distribution of the time from hospitalization to outcome (discharge/death) among COVID-19 cases in the provinces other than Hubei as of February 23. As a sensitivity analysis, we also considered the 80^th^ and 50^th^ percentiles of this distribution, corresponding to 14 days and 10 days, respectively. As mentioned above, patients were discharged only after testing negative by nucleic acid detection tests on two consecutive days^9^. Hence, we assumed that these patients had biologically recovered three days prior to the reported date of discharge, accounting for one additional day for delay of laboratory confirmation and official reporting. Accordingly, for the discharged patients, their time from hospitalization to discharge was cut down by three days when estimating the 90^th^, 80^th^ and 50^th^ percentiles of the distribution of the time from hospitalization to outcome.

All deaths occurred among critical cases, as reported by China CDC^5^. Separately for mainland China, Hubei Province, and the provinces outside Hubei, we further estimated CFRs among severe and critical patients by dividing the above derived CFR by proportions of severe and critical patients among all reported COVID-19 cases. We used the average of daily proportions among COVID-19 cases who were still in hospitals on each day other than the clinical severity on admission, which were obtained from the aggregated dataset and showed very stable (Table S2, and Figure S2-3 in Appendix). Only Guangdong Province officially reported aggregated data on mild-, moderate-, severe- and critical patients. And thus, for the provinces outside Hubei, we additionally estimated the CFR by these clinical categories using the corresponding proportions in Guangdong Province. Statistical analyses were performed with R (version 3.6.0).

### Ethics

The study was approved by the Institutional review board from School of Public Health, Fudan University (IRB#2020-02-0802). All data were collected from publicly available sources and did not contain any personal information.

## Results

As of February 23, 2020, a total of 77,150 laboratory-confirmed cases with 2,592 deaths, 24,711 discharged and 49,847 patients who were still hospitalized were reported in mainland China (see Table S2 for details of each province). Of these, provinces outside Hubei accounted for 12,863 (16.7%, 12,863/77,150) laboratory-confirmed cases including 97 deaths (3.7%, 97/2,592), 7,973 (32.3%, 7,973/24,711) discharged cases and 4,793 (9.6%, 4,793/49,847) patients who were still hospitalized. We collected individual information from publicly available official sources on 9,651 laboratory-confirmed cases detected outside Hubei by February 23, accounting for 75.0% (9,651/12,863) of total cases reported, 63.9% (62/97) of deceased patients, 18.2% (1,449/7,973) of recovered patients. Of 9,651 cases, unresolved patients accounted for 84.3% (8,140/9,651) (Table 1). See Figure S4 for the epidemic curve of cases with available individual information.

**Table 1.**
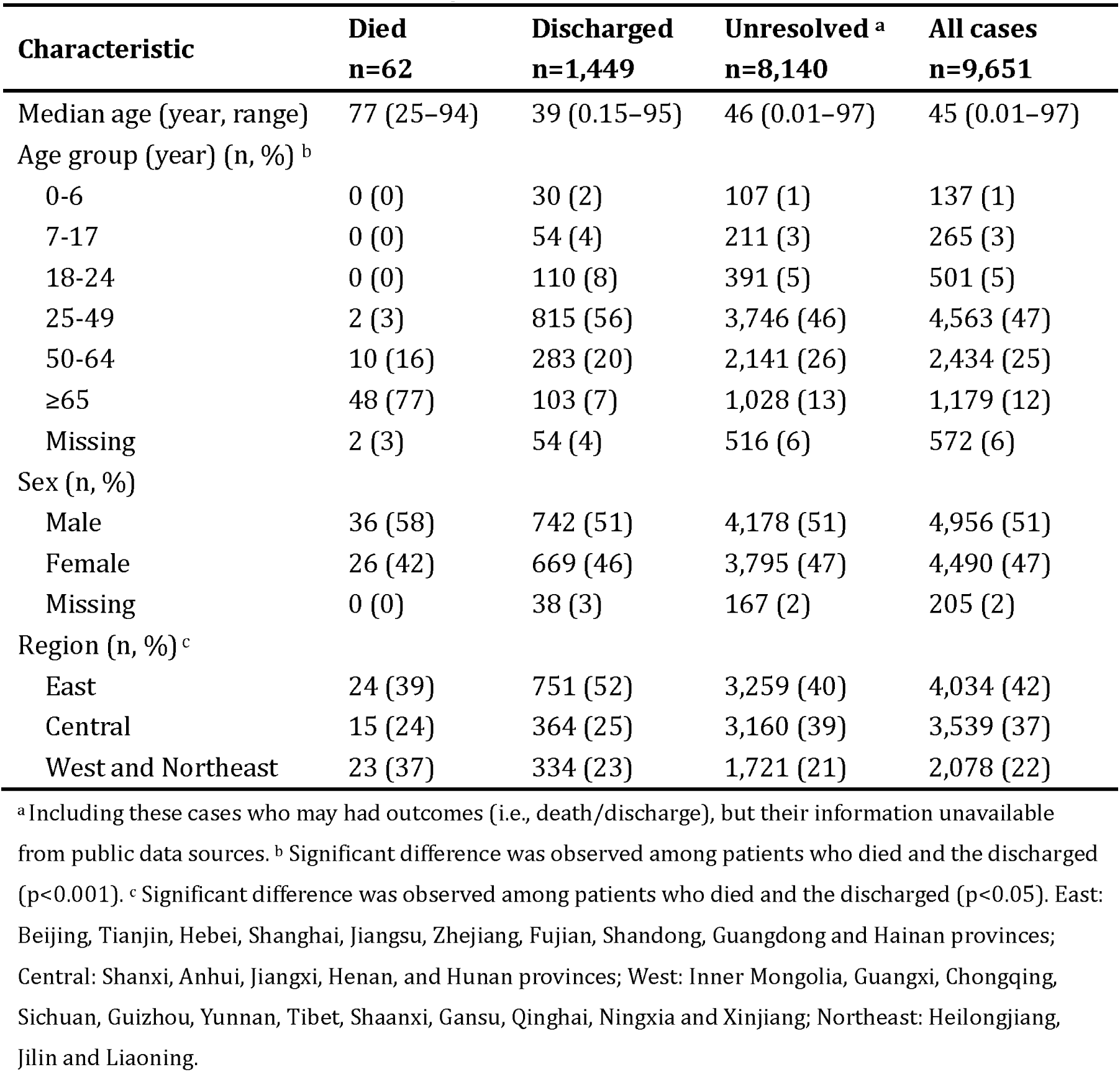
Demographical characteristics of COVID-19 cases outside Hubei province in mainland China, as of February 23, 2020

The median age of the cases outside Hubei was 45 years (range, four days-97 years), and 51% (4,956/9,651) were male. Those who died were significantly older than discharged cases (median age: 77 vs 39 years, p<0.001). 77% (48/62) of deaths occurred in the older adults aged 65 years or above, and 58% (36/62) were male. (Table 1) Multivariate logistic analysis revealed that increasing age, being male, and living in less developed economic regions (e.g. Central region or West and Northeast region) were risk factors for mortality (Table 2). The univariate logistic analysis is shown in Appendix Table S5.

**Table 2.** Risk factors associated with fatal outcome among COVID-19 patients

| Variables | OR (95%CI) | Z-value | P-value |
| --- | --- | --- | --- |
| Age, per year increase | 1.18 (1.14-1.22) | 10.2 | <0.001 |
| Sex |  |  |  |
| Female | ref | / | / |
| Male | 2.02 (1.02-4.03) | 2.00 | 0.045 |
| Unknown | 0 (0-Inf) | -0.01 | 0.990 |
| Economic regions <sup>a</sup> |  |  |  |
| East | ref | / | / |
| Central | 3.45 (1.32-9.03) | 2.53 | 0.012 |
| West and Northeast | 3.93 (1.74-8.85) | 3.30 | <0.001 |
| Time from symptom onset to first healthcare consultation |  |  |  |
| ≤2 days | ref | / | / |
| >2 days | 1.11 (0.33-3.71) | 0.17 | 0.863 |
| Unknown | 0.40 (0.14-1.14) | -1.72 | 0.086 |
| Time from symptom onset to hospital admission |  |  |  |
| ≤3 days | ref | / | / |
| >3 days | 0.65 (0.20-2.11) | -0.72 | 0.471 |
| Unknown | 0.55 (0.18-1.69) | -1.04 | 0.298 |
| Time from symptom onset to laboratory confirmation |  |  |  |
| ≤6 days | ref | / | / |
| >6 days | 1.41 (0.42-4.74) | 0.56 | 0.575 |
| Unknown | 1.41 (0.44-4.55) | 0.58 | 0.561 |

<sup>a</sup> East: Beijing, Tianjin, Hebei, Shanghai, Jiangsu, Zhejiang, Fujian, Shandong, Guangdong and Hainan provinces; Central: Shanxi, Anhui, Jiangxi, Henan, and Hunan provinces; West: Inner Mongolia, Guangxi, Chongqing, Sichuan, Guizhou, Yunnan, Tibet, Shaanxi, Gansu, Qinghai, Ningxia and Xinjiang; Northeast: Heilongjiang, Jilin and Liaoning. /not applicable.

The intervals from symptom onset to first healthcare consultation, from symptom onset to hospitalization, and from symptom onset to laboratory confirmation were consistently longer for deceased patients than for those who recovered. Overall, the time interval from symptom onset to death was estimated to be 12.9 days (95%CI: 2.2 to 40.2), and from symptom onset to discharge was 16.7 days (95%CI: 8.6 to 28.9). (Table 3)

**Table 3.** Key time to event intervals of COVID-19 patients outside Hubei province in mainland China, as of February 23, 2020 (mean, 95%CI)

| Key time to event interval | Died<br>n=62 | Discharged<br>n=1,449 | Unresolved <sup>a</sup><br>n=8,140 |
| --- | --- | --- | --- |
| Time from symptom onset to first healthcare consultation (days) | n=27 | n=493 | n=2,838 |
| Estimates from empirical data | 3.0 (0.5, 11.1) | 1.0 (0.5, 11.0) | 2.0 (0.5, 11.0) |
| Estimates by fitting | 2.0 (0.2, 18.2) | 1.6 (0.2, 13.2) | 1.6 (0.2, 12.5) |
| Time from symptom onset to hospital admission (days) | n=36 | n=572 | n=2,148 |
| Estimates from empirical data | 4.0 (0.5, 13.2) | 3.0 (0.5, 12.0) | 3.0 (0.5, 12.0) |
| Estimates by fitting | 3.4 (0.1, 17.0) | 3.1 (0.2, 13.1) | 2.2 (0.3, 18.0) |
| Time from symptom onset to laboratory confirmation (days) | n=35 | n=654 | n=4,955 |
| Estimates from empirical data | 6.0 (0.5, 16.0) | 5.0 (0.7, 15.0) | 5.0 (0.5, 16.0) |
| Estimates by fitting | 5.6 (0.6, 17.2) | 5.1 (0.6, 15.3) | 5.2 (0.6, 15.8) |
| Time from symptom onset to death/discharge (days) | n=41 | n=769 | / |
| Estimates from empirical data | 13.0 (3.0, 39.0) | 17.0 (8.0, 29.0) | / |
| Estimates by fitting | 12.9 (2.2, 40.2) | 16.7 (8.6, 28.9) | / |
| Time from hospital admission to death/discharge (days) | n=49 | n=980 | / |
| Estimates from empirical data | 8.0 (0.6, 35.4) | 13.0 (6.0, 24.5) | / |
| Estimates by fitting | 8.0 (0.8, 30.2) | 13.1 (6.0, 24.2) | / |

<sup>a</sup> Including these cases who may had outcomes (i.e., death/discharge), but their information unavailable from publicly data sources; / not applicable.

Based on the total patients reported to the surveillance system, the CFR estimated by Garske’s method^11^ and survival analyses were all higher than the crude CFR (Table 4 and Appendix Table S6). The CFR estimated by Garske’s method^11^ was 4.52% (95%CI: 4.47% to 4.67%) in mainland China, with highest estimate in Wuhan (6.19%, 95%CI: 6.12% to 6.41%), and lowest in the provinces outside Hubei (0.89%, 95%CI: 0.83% to 1.06%). The CFR estimated by survival analyses was 1.24% (95%CI: 1.24% to 1.24%) among all cases, and 11.21% (95%CI: 11.21% to 11.21%) among severe and critical patients outside Hubei. There was no difference in both overall CFR and that among severe and critical patients outside Hubei estimated by survival analyses or Garske’s method (p>0.05) (Table 4). In sensitivity analyses, we excluded all cases hospitalized for longer than 14 and 10 days, and the estimates were all consistent with those of the baseline analysis, in which we excluded cases with hospitalizations longer than 17 days (p>0.05) (Appendix, Figure S5-6).

**Table 4.** Fatality risk of COVID-19 among all reported cases, and among severe and critical cases ^a^

|  | Number of cases |  | Fatality risk among all reported cases<br>(%, 95%CI) |  |  | Fatality risk among severe and critical patients<br>(%, 95%CI) <sup>b</sup> |  |  |
| --- | --- | --- | --- | --- | --- | --- | --- | --- |
|  | Death | Total cases reported | Crude | Estimated using Garske's method<sup>11</sup> | Estimated by survival analyses | Crude | Estimated using Garske's method<sup>11</sup> | Estimated by survival analyses |
| Mainland China | 2,592 | 77,150 | 3.36<br>(3.23, 3.49) | 4.52<br>(4.47, 4.67) | / | 17.97<br>(17.30, 18.66) | 24.18<br>(23.91, 24.96) | / |
| Hubei province | 2,495 | 64,287 | 3.88<br>(3.73, 4.03) | 5.37<br>(5.31, 5.55) | / | 18.57<br>(17.86, 19.30) | 25.71<br>(25.42, 26.54) | / |
| Wuhan | 1,987 | 46,607 | 4.26<br>(4.08, 4.45) | 6.19<br>(6.12, 6.41) | / | 18.54<br>(17.75, 19.35) | 26.93<br>(26.61, 27.88) | / |
| Outside Wuhan | 508 | 17,680 | 2.87<br>(2.63, 3.13) | 3.54<br>(3.44, 3.81) | / | 23.36<br>(21.4, 25.45) | 28.75<br>(28.00, 30.99) | / |
| Outside Hubei province | 97 | 12,863 | 0.75<br>(0.62, 0.92) | 0.89<br>(0.83, 1.06) | 1.24<br>(1.24, 1.24) | 6.79<br>(5.57, 8.28) | 8.02<br>(7.48, 9.55) | 11.21<br>(11.21, 11.21) |

<sup>a</sup> crude fatality risk was calculated as the cumulative number of deaths divided by the cumulative number of laboratory-confirmed cases.
/not estimated using survival analyses due to limited individual data.
<sup>b</sup> estimated using the proportion of severe and critical patients among reported cases in corresponding areas.

In the provinces other than Hubei, the CFR increased with age, with highest estimates among patients aged ≥70 years (Figure 1 panel B). The fatality risk among critical patients was 23.8-33.3%, which was 2-fold higher than that among severe and critical patients, and 24-fold higher than that among moderate, severe and critical patients (Figure 1 panel B). The CFR among all cases estimated by survival analyses declined rapidly from 8% on January 25 to around 1% on January 28, and remained at 1.2-1.5% afterwards. Patterns were similar for estimates using Garske’s method (Figure 2).

**Figure 1.**
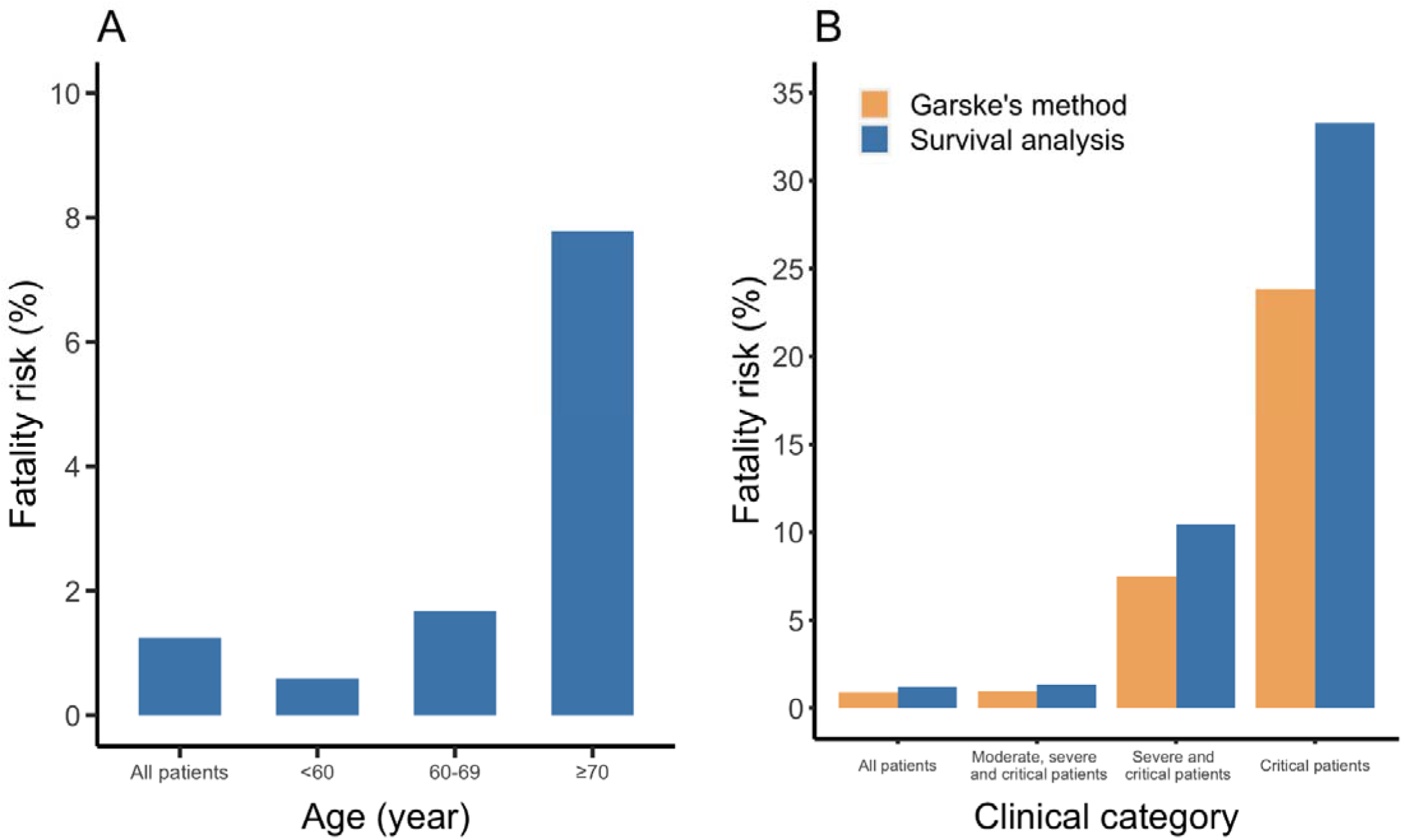
Case-fatality risk (mean) outside Hubei province in mainland China. A: by age group; B: by clinical categories (All patients includes mild, moderate, severe and critical patients). 95%CI was narrow and thus not presented here.

**Figure 2.**
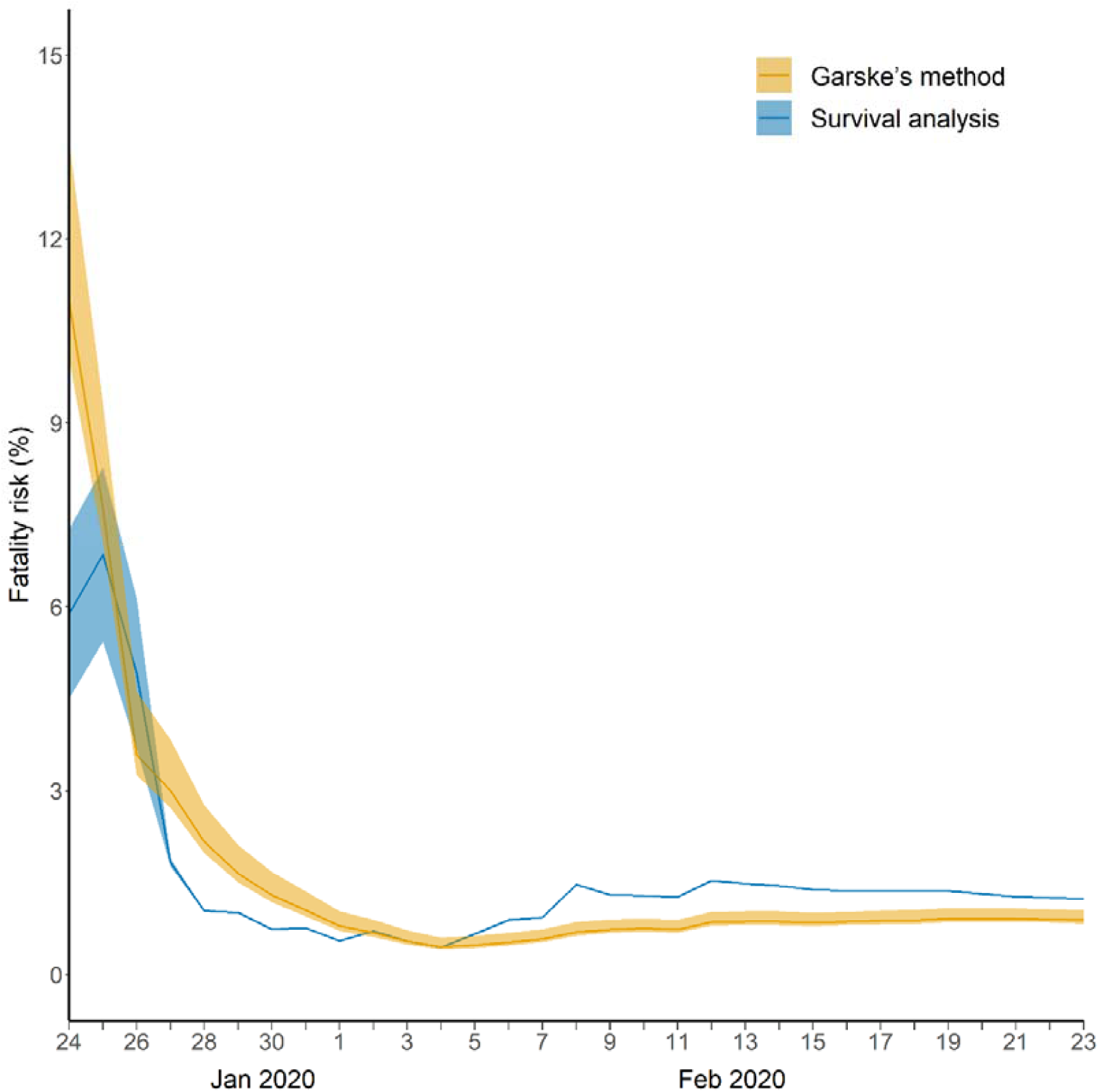
Case-fatality risk over time outside Hubei province in mainland China (%) (mean, 95%CI).

## Discussion

We have shown that the fatality risk among detected cases was 0.89-1.24% in the provinces outside Hubei in mainland China and increased with clinical severity. Further, the CFR was estimated at 8.02-11.21% among severe and critical patients. Estimates accounting for right-censoring of unresolved cases were higher than crude estimates. Male patients, older age, and less developed regions were factors associated with a higher CFR. These estimates could represent the most accurate estimates of CFR in China so far.

Our study is strengthened by accounting for unknown outcomes among patients who were still in hospital at time of data cutoff. We used Garske’s method developed for pandemic influenza A/H1N1 in 2009^11^, as well as survival analyses, both of which consider censoring. Compared to the crude CFR, Garske’s method improved estimates by adjusting for the cumulative density of intervals from hospital admission to death/discharge. Survival analysis was a useful tool for comparison as it relied on a very large individual dataset comprising a total of 9,651 reported cases. Availability of individual data enabled us to explore mortality risk factors and estimate CFR by age group. Estimates have not been reported previously based on such a large sample size and a competing risk model of survival analysis with 90^th^ quantile truncation. There was no difference in CFR estimates by the two methods, which lends support to our estimates.

Our study has some limitations. First, in the individual dataset, the clinical profile of patients was not available. Hence, we could not provide direct estimates of fatality risk stratified by clinical categories using survival analysis. Instead, we divided the estimated CFR among all cases by the proportions of different clinical categories obtained from the aggregated dataset. This is a reasonable approach method because all deaths occurred among critical cases ^5^.

Second, the analyzed individual records were retrieved from publicly available official sources, ensuring accuracy and reliability of information. However, we were only able to collect few individual records in Hubei because they did not release complete individual information. And thus, we were unable to estimate CFR in Hubei using survival analyses. Moreover, assessment of clinical severity in Hubei, especially in the epicenter of the outbreak in Wuhan, is challenging because disease severity may be increased by bottlenecks in local healthcare capacity, as COVID-19 cases surged. In addition, case surveillance and clinical management were biased towards severe cases in Hubei, especially in the early phase of the epidemic. Our estimates of the CFR in Hubei and Wuhan using Garske’s method^11^ should be viewed cautiously as the sensitivity of surveillance of both deaths and cases remains unclear.

Our study only addresses CFR among detected cases. The level of ascertainment of mild cases remains unclear. More estimates that include fatality risk among syndromic patients and asymptomatically infected individuals can only be available through enhanced routine surveillance, such as increased testing of patients with influenza-like-illnesses, and by analysis of future sero-epidemiological studies.

Our CFR estimates of 0.89-1.24% among detected COVID-19 patients outside Hubei province are higher than the crude CFRs reported by WHO and China CDC, which is 0.4-0.7%^5,14^. It is expected that the crude CFR obtained by dividing the cumulative number of reported deaths by the cumulative number of reported cases is an underestimate due to the inevitable delay between symptom onset and death. Our findings reveal that older individuals and male patients experience higher fatality risk, which is consistent with the WHO report ^14^.

Additionally, WHO reported that patients with underlying conditions had much higher fatality rates^14^. Our study was unable to address the relative risk of fatal outcome among patients with underlying diseases compared to healthy people, because limited information on underlying conditions was available from publicly available data sources.

A clinical study conducted in Wuhan showed that 4.3% of hospitalized patients died^15^. Another study relying on patients from 552 hospitals across 30 provinces, found that 1.4% of patients died ^16^, in which less study participants (28%) were from Wuhan. The estimates of these clinical studies would be the lower bounds for the CFR since separately 62% and 94% of patients were still in hospitals. The fatality risk from these clinical studies is higher than our CFR estimates, probably due to shortage of health services in Wuhan, e.g., advanced health care facilities for critically ill patients as extracorporeal membrane oxygenation.

Our CFR estimates outside Hubei province indicate that the severity of SARS-CoV-2 is lower than that of other diseases caused by zoonotic coronaviruses, including Middle East respiratory syndrome (MERS), which had an estimated CFR of 34.4% globally^17^, and severe acute respiratory syndrome (SARS) with an estimated CFR of 10.9% across the world and 7% in mainland China^18^. In contrast, the CFR of SARS-CoV is in the same order of magnitude as that of pandemic 2009 influenza A(H1N1) virus hospitalizations, which has an estimated CFR of 1.4% among hospitalized patients in Asia^19^. In the long run, depending on how much of the severity pyramid of SARS-CoV2 is captured in our data, the absolute severity of SARS-CoV2 may prove to be similar, to somewhat more severe, than the 2009 influenza pandemic, albeit with a different age profile. Comparison of clinical data from China and other countries will prove useful to settle this question.

Outside Hubei province, close contacts of laboratory-confirmed cases were kept in quarantine for 14 days, and local hospitals tested patients with respiratory symptoms (e.g., fever and cough) and epidemiological links to Hubei province or other cases. This strategy would have enabled detection of many mild cases. However, a small number of mild cases were captured. In our aggregated dataset for Guangdong province for instance, only 8% of reported cases were mild, while the majority (83%) of reported cases had moderate disease severity with presence of pneumonia. And thus, our CFR estimates could approximately represent the fatality risk among laboratory-confirmed COVID-19 cases with chest x-ray confirmed pneumonia. Even though clinical information for these patients was not available from publicly available sources, we believe that our CFR estimates could be viewed as the fatality risk among hospitalized COVID-19 cases. Chest x-ray confirmed pneumonia is a threshold for hospital admissions in in China. This may vary among countries due to different clinical practices and health service capacity.

Clinical studies have reported a higher proportion of severe patients among older age group (29% vs. 14%)^16^. No specialized treatment for COVID-19 patients has been identified, and the mainstay clinical management has been supportive care. For non-critically ill patients, close follow-up is likely to be sufficient to manage the disease. But critically ill patients were more likely to develop ARDS and require ICU admission^20^. That could explain our findings that severe patients had a higher fatality risk. The high observed CFRs of COVID-19 in older adults is consistent with the age profile of MERS, SARS, pandemic H1N1 2009, and seasonal influenza.^19^

Compared to the Eastern region, cases detected in the less developed Central region had a 2.45-fold higher risk of death, and those in West and Northeast region had a 2.93-fold higher risk. It is important to note that those variations in CFR do not reflect underlying differences in clinical disease severity. CFR will vary regionally depending on the sensitivity of surveillance systems to detect cases at different levels of the severity pyramid and clinical care offered to severe and critical patients. More attention should be paid to less developed settings with limited health services like Iran, which reports a larger ratio of deaths to cases than other countries^2^.

Notably, the definition of suspected cases eligible for laboratory testing used in China shifted from a narrow clinical criteria based on three symptoms early in the outbreak (fever; radiographic findings of pneumonia; normal or reduced white blood cell count, or reduced lymphocyte count at early onset of symptoms), to a broader criteria including any two of three symptoms by January 27. This would bias our sample towards more clinically severe cases before January 27. In addition to improvement in therapeutic capacity, the shift in surveillance definition could partially explain the declining trend of CFR from 8% to around 1% at the end of January, which remained stable afterwards. Accordingly, our CFR estimate for February could provide a true picture of the severity of laboratory-confirmed cases of COVID-19.

In conclusion, our estimates of CFR among laboratory-confirmed cases suggest that COVID-19 is not as severe as SARS and MERS, but similar to that of pandemic 2009 H1N1 among hospitalized patients. The fatality risk of COVID-19 cases is higher in male, and increases with age, particularly in adults aged 70 years and above. Our findings can inform the severity assessment and response to the on-going COVID-19 outbreak, and assist preparations for a global epidemic of COVID-19.

## Data Availability

no additional data available.

## Contributors

H.Y. conceived, designed and supervised the study. W.W., J.L., Y.C., H.Y., Y.Z., Q.Q., H.G., Xiang.W., L.W. and K.S. participated in data collection. X.D., J.Y., X.W., JX.Z., Z.C, J.Z., and Y.W. analyzed the data, and prepared the figures. J.Y. prepared the first draft of the manuscript. X.D., P.W., M.A., B.C., C.V., and H.Y. commented on the data and its interpretation, revised the content critically. All authors contributed to review and revision and approved the final manuscript as submitted and agree to be accountable for all aspects of the work.

## Disclaimer

The findings and conclusions in this study are those of the authors and do not necessarily represent the official position of the National Institutes of Health or U.S. Department of Health and Human Services.

## Acknowledgments

We thank Xin Chen, Jiaxian Chen, and Sihong Zhao, from School of Public Health, Fudan University, and Yuheng Feng from School of Basic Medical, Sciences, Fudan University for providing assistance with data collection.

## Notes

### Competing Interest Statement

H.Y. has received research funding from Sanofi Pasteur, GlaxoSmithKline, Yichang HEC Changjiang Pharmaceutical Company, and Shanghai Roche Pharmaceutical Company. BJC has received honoraria from Roche and Sanofi. None of those research funding is related to COVID-19. All other authors report no competing interests.

### Funding Statement

H.Y. acknowledges financial support from the National Science Fund for Distinguished Young Scholars (No. 81525023), Key Emergency Project of Shanghai Science and Technology Committee (No. 20411950100), National Science and Technology Major Project of China (No. 2018ZX10201001-010, No. 2018ZX10713001-007, No. 2017ZX10103009-005).

## Reference

1. National Health Commission of the People’s Republic of China. Update on COVID-19 as of 24:00 on March 3, 2020. http://www.nhc.gov.cn/xcs/yqtb/202003/7a5f57b3f1b94954b1fc25f81dacc874.shtml (accessed March 4 2020).

2. World Health Organization. Coronavirus disease 2019 (COVID-19) Situation Report – 343 2020. https://www.who.int/docs/default-source/coronaviruse/situation-reports/20200303-sitrep-43-covid-19.pdf?sfvrsn=2c21c09c_2 (accessed March 4 2020).

3. Wu P, Hao X, Lau EHY, et al. Real-time tentative assessment of the epidemiological characteristics of novel coronavirus infections in Wuhan, China, as at 22 January 2020. Euro Surveill 2020.

4. WHO Collaborating Centre for Infectious Disease Modelling and Imperial College London. Dorigatti I, Okell L, Cori A, Imai N, Baguelin M, Bhatia S, et al. Report 4: Severity of 2019-novel coronavirus (nCoV). https://www.imperial.ac.uk/mrc-global-infectious-disease-analysis/news--wuhan-coronavirus/. (accessed Feb 25 2020).

5. The Novel Coronavirus Pneumonia Emergency Response Epidemiology Team. The Epidemiological Characteristics of an Outbreak of 2019 Novel Coronavirus Diseases (COVID-19) — China, 2020. China CDC Weekly 2020; 2(8):113–22.

6. Lipsitch M, Donnelly CA, Fraser C, et al. Potential Biases in Estimating Absolute and Relative Case-Fatality Risks during Outbreaks. PLoS Negl Trop Dis 2015; 9(7): e0003846.

7. Prevention CCfDCa. Epidemic update and risk assessment of 2019 Novel Coronavirus. 2020. http://www.chinacdc.cn/yyrdgz/202001/P020200128523354919292.pdf (accessed Jan 31 2020).

8. National Health Commission of China. The diagnosis and treatment scheme of novel coronavirus diseases 2019 (Trial version 5th). http://www.gov.cn/zhengce/zhengceku/2020-02/05/content_5474791.htm. (accessed Feb 25 2020).

9. National Health Commission of China. The diagnosis and treatment scheme of novel coronavirus diseases 2019 (Trial version 6th). http://www.gov.cn/zhengce/zhengceku/2020-02/19/content_5480948.htm. (accessed Feb 25 2020).

10. National Bureau of Statistics of China. China Statistical Yearbook 2019. http://www.stats.gov.cn/tjsj/ndsj/2019/indexch.htm. (accessed Feb 25 2020)

11. Garske T, Legrand J, Donnelly CA, et al. Assessing the severity of the novel influenza A/H1N1 pandemic. BMJ 2009; 339: b2840.

12. Jewell NP, Lei X, Ghani AC, et al. Non-parametric estimation of the case fatality ratio with competing risks data: an application to Severe Acute Respiratory Syndrome (SARS). Stat Med 2007; 26: 1982–98.

13. Rubin DB. Multiple imputation for nonresponse in surveys. New York, NY: J Wiley and Sons, 1987.

14. World Health Organization. Report of the WHO-China Joint Mission on Coronavirus Disease 2019 (COVID-19). https://www.who.int/docs/default-source/coronaviruse/who-china-joint-mission-on-covid-19-final-report.pdf. (accessed on Feb 29 2020).

15. Wang D, Hu B, Hu C, et al. Clinical Characteristics of 138 Hospitalized Patients With 2019 Novel Coronavirus-Infected Pneumonia in Wuhan, China. JAMA 2020: 10.1001/jama.2020.1585.

16. Guan W-J, Ni Z-Y, Hu Y, et al. Clinical Characteristics of Coronavirus Disease 2019 in China. The New England journal of medicine 2020: 10.1056/NEJMoa2002032.

17. World Health Organization. Middle East respiratory syndrome coronavirus (MERS-CoV) – The Kingdom of Saudi Arabia. https://www.who.int/csr/don/18-december-2019-mers-saudi-arabia/en/. (accessed on Feb 29 2020).

18. World Health Organization.Summary of probable SARS cases with onset of illness from 1 November 2002 to 31 July 2003. https://www.who.int/csr/sars/country/table2003_09_23/en/. (accessed on Feb 29 2020).

19. Wong JY, Kelly H, Cheung C-MM, et al. Hospitalization Fatality Risk of Influenza A(H1N1)pdm09: A Systematic Review and Meta-Analysis. Am J Epidemiol 2015; 182(4): 294–301.

20. Yang X, Yu Y, Xu J, et al. Clinical course and outcomes of critically ill patients with SARS-CoV-2 pneumonia in Wuhan, China: a single-centered, retrospective, observational study. Lancet Respir Med 2020: S2213-600(20)30079-5.

